# An assessment of the prevalence and risk factors of hypertensive crisis in patients who visit the Emergency Outpatient Department (EOPD) at Adama Hospital Medical College, Adama, Oromia, Ethiopia: A 6-month prospective study

**DOI:** 10.1101/2023.09.01.23294941

**Authors:** Abel Tezera Abebe, Yabets Tesfaye Kebede, Bekri Delil Mohammed

## Abstract

**Background:** Over 1 billion people worldwide suffer from the common chronic medical condition of hypertension. A hypertensive crisis occurs when blood pressure exceeds 180/110 mmHg. Depending on whether the target organ is harmed, the situation may be presented as a hypertensive emergency or urgency.

**Objective:** To assess the prevalence and risk factors of patients with hypertensive crises who visited the Emergency Outpatient Department (EOPD) at Adama Hospital Medical College in Adama, Oromia, Ethiopia, between January 01 and August 31, 2021 G.C.

**Methodology:** A cross sectional, prospective study on hypertensive crisis was conducted at Adama Hospital Medical College from January 01 to August 31, 2021 G.C. The data was collected using a standardized questionnaire, validated for completeness, and analyzed using SPSS. The findings were tabulated, and conclusions and recommendations were conveyed.

**Result:** A total of 444 individuals with hypertension in crisis were identified. Of these, 56.8% were men, resulting in a M:F ratio of 1.31:1. Those between the ages of 66 and 75 were the most affected. At presentation, 91.0% of the study participants were known hypertensive patients. Of the known hypertensive patients, the majority (34.9%) were known to have been hypertensive for 5 to 10 years. Of the known hypertensive patients, 48.6% were found to be adherent. Hypertensive urgency was discovered to be far more common than hypertensive emergencies (63.5% vs. 36.5%). Headache was the most common presenting symptom, and most of the patients (36.5%) presented to the health setup in less than 24 hours. The main risk variables identified were drug discontinuation, family history of hypertension, salt consumption, and alcohol usage. The main excuse for lack of adherence was the cost of the medications. More than half of the patients do not have any additional comorbidities, and of those who do, diabetes mellitus is the most prevalent. A stroke was identified as a major complication.

**Conclusions and recommendations:** Hypertensive crises are one of the most prevalent reasons for EOPD admission and are linked to significant consequences. At presentation, most of the study subjects were known hypertension patients. Diabetes mellitus was discovered to be a comorbid condition in one quarter of them. Although more than half of the patients have improved, the death rate remains high. Infrastructure and capacity building to provide hospitals with the requisite baseline investigations are among the government’s recommendations. Health practitioners are expected to make improvements, such as educating the public about the need for lifestyle changes and evaluating and managing any hypertension problems.

## INTRODUCTION

### Background information

One of the primary causes of the global disease burden is hypertension. Over a billion people worldwide have high blood pressure, which is considered the cause of 9.4 million annual deaths. Cardiovascular diseases like coronary heart disease, congestive heart failure, ischemic and hemorrhagic stroke, renal failure, and peripheral arterial disease are all twice as likely to occur in people with hypertension. Although antihypertensive medications lower the risk of cardiovascular and renal disease, a sizable portion of hypertension sufferers go untreated or receive insufficient care.

Given that hypertension is a major global public health concern, its prevention, detection, treatment, and control should be given top priority. However, the majority of countries in Sub-Saharan Africa (SSA) are still battling infectious diseases like HIV, malaria, and tuberculosis, and the majority of their governments lack adequate funding for health care (1). A rise in the prevalence of hypertension in this area is therefore likely to have negative effects.

Previously, the Joint National Committee classified hypertension into four stages: normal (less than 120/80 mmHg), pre-HTN (between 120 and 139/80 and 89 mmHg), stage I (between 140 and 159/90 and 99 mmHg), and stage II (above 160/100 mmHg). The new ACC/AHA guidelines, the first comprehensive set since 2003, changed the definition of high blood pressure to account for complications that can arise at lower readings and to enable earlier intervention. BP categories according to the new ACC/AHA guideline are normal (below 120/80 mmHg), elevated (SBP between 120 and 129 and DBP below 80 mmHg), stage 1 (SBP between 130 and 139 and DBP between 80 and 89 mmHg), and stage 2 (SBP at least 140 or DBP at least 90 mmHg).

The Eighth Joint National Committee (2) categorizes blood pressure elevations as either “hypertensive urgencies,” which include asymptomatic severe hypertension without target organ damage, or “hypertensive emergencies,” which include acute, life-threatening conditions associated with marked increases in blood pressure, typically 180/120 mmHg. It is crucial to distinguish between hypertensive emergencies and urgency when developing a treatment plan. The former seeks to lower blood pressure gradually over several hours to days to 160/100 mmHg, whereas the latter seeks to lower blood pressure right away (although not necessarily to normal ranges) in order to prevent or limit the deterioration of the target organ. (3, 4)

It should be noted that acute end-organ damage rather than a specific blood pressure level, distinguishes hypertensive urgency from an emergency. A hypertensive emergency is a clinical diagnosis, and the clinical condition of the patient is more significant than the blood pressure’s absolute value. (3)

The phrase “hypertensive emergency or crisis” has replaced the term “malignant hypertension” in national and international blood pressure control recommendations. Malignant hypertension was once used to describe a condition characterized by elevated blood pressure combined with encephalopathy or acute nephropathy. (2)

### Statement of the Problem

The high incidence of hypertension and the risk of cardiovascular disease it carries make it a major public health concern worldwide. (5) The global prevalence of hypertension is expected to exceed 29% by 2025. In Sub-Saharan Africa (SSA), approximately 75 million people (roughly twice the population of California) have hypertension, with a projected 125.5 million people affected by 2025 (20), and more than 80% of deaths from hypertension and associated cardiovascular diseases occur in low- and middle-income countries. (6)

In the industrialized world, hypertension is becoming a more common health condition due to an extended lifespan and the prevalence of risk factors like obesity, inactivity, and poor diet. The frequency is already comparable to that seen in wealthy nations in many emerging nations, especially in urban cultures. (7)

Uncontrolled hypertension continues to be a major contributor to cardiovascular disease despite the availability of effective treatments. (8) As a result, when systolic blood pressure/diastolic blood pressure (SBP/DBP) is greater than 180/120 mmHg, suboptimal BP control might result in a hypertensive crisis. (9)

The treatment of hypertension has been linked to a 40% reduction in the risk of stroke and a 15% reduction in the risk of myocardial infarction. Even though HTN medication has been demonstrated to prevent CVD and to extend and improve life, hypertension is still poorly treated around the world. (7) Furthermore, obesity, diabetes, hyperlipidemia, and tobacco use are all common co-occurring cardiovascular risk factors with hypertension, all of which raise the cardiovascular risk related to hypertension. Insufficient attention is given to these coexisting risk factors in hypertensive patients around the world, which has a high morbidity and mortality rate.

Over the past 40 years, hypertensive crises have become more common despite the development of increasingly effective antihypertensive medications. (10) Given that most hypertensive emergencies involve people with chronic hypertension, this may be related to a lack of awareness on the part of society. This is mainly because people don’t take their medications as prescribed; they get high on stimulants like cocaine; and they experience withdrawal symptoms from antihypertensive medications like clonidine and beta blockers.

Although it is well known that treating hypertensive emergencies quickly is necessary to avoid permanent or worsened target organ damage, (8) this does not seem to apply in our setting because of a number of factors, including gaps in patients’ full clinical history, inadequate laboratory and radiographic equipment, and a lack of blood pressure cuffs that are the right size, which is crucial because using a cuff that is too small for the arm size has been shown to artificially elevate BP.

There is a lack of data on the types of hypertensive emergencies seen in Sub-Saharan Africa, as well as the symptoms and outcomes. Most notably in Ethiopia, little is known about the prevalence, risk factors, and prognosis of hypertensive emergencies in East Africa. Data on these tendencies is critical for increasing physician awareness. Raising awareness of the community’s risk factors will aid in lowering morbidity and mortality rates among these hypertensive patients.

### Significance of the study

According to numerous studies, the majority of people with hypertensive crises who stopped taking their medication for a significant period of time did so because they felt better. This result demonstrates the significance of effective hypertension control in reducing high blood pressure-related complications.

Health professionals in our situation would be better able to assess the severity of the issue and develop effective solutions if they were aware of the risk factors for hypertensive emergencies. It will serve as a starting point for additional research in the field by a number of governmental and non-governmental organizations involved in the management of non-communicable diseases like hypertension. The study will offer a helpful manual for people to follow in order to lessen the financial, social, psychological, and physical effects of hypertension.

This research could therefore aid in improving the management and rehabilitation of hypertensive patients as well as the prevention of the emergence of chronic hypertension complications, primarily brought on by patients’ poor adherence to hypertensive regimens and insufficient care and follow-up for the condition.

## OBJECTIVES

### General objectives

► To assess the prevalence and risk factors of hypertensive Crisis among patients who visit the Emergency Outpatient Department (EOPD) at Adama Hospital Medical College, 2021 G.C

### Specific objectives

► To assess the socio-demographic characteristics of patients with hypertensive crisis.
► To assess the risk factors for hypertensive crisis among patients who visited (EOPD) at Adama Hospital Medical College (AHMC), 2021 G.C.
► To assess the complications of hypertensive crisis among patients who visited (EOPD) at Adama Hospital Medical College (AHMC), 2021 G.C.

## METHODOLOGY

### Study area

Adama Hospital Medical College (AHMC) was previously known by the names of Hayile Mariam Mamo Memorial Hospital and Adama Referral Public Hospital at different times. It is one of the first medical hospitals situated in Adama town, located in the Oromia Region, 100 kilometers (about 62.14 miles) southeast of Addis Abeba, Ethiopia. The hospital was inaugurated by missionaries from abroad in 1938 E.C. and was among the first non-governmental hospitals in the nation.

The hospital was upgraded to a medical college in 2003 EC because of its location, patient load, and staff capacity. The hospital is serving a catchment population of more than 6 million from five regions (Oromia, Amhara, Afar, Somali, and Dire-Dewa). The hospital has 232 beds and serves 1,000 patients daily through six medical case teams (OPD) and various specialty clinics.

### Study period

- The study period was from January 01 to August 31, 2021 G.C.

### Study design

- A prospective institution-based cross-sectional study was conducted.

### Source Population

- The source populations of the study were patients who presented with Hypertensive Crisis to EOPD in Adama Hospital Medical College (AHMC) from January 01 to August 31, 2021 G.C.

### Study population

- The study population will consist of any selected patient with hypertension who meets the study’s inclusion criteria.

### Inclusion criteria

- All adult patients over the age of 18 who presented with hypertensive crisis to the EOPD at Adama Hospital Medical College during the study period met the inclusion criteria.

### Exclusion criteria

- Any patient who is younger than 18 years old.
- Incomplete data or those who self-discharged after being seen at the EOPD were excluded from the study.

### Sample size and sample technique

- Sampling was not used because all cases of patients diagnosed with hypertensive crisis after presenting to the hospital during the previously mentioned study period were included.

### Dependent Variables

- prevalence of hypertensive crisis

### Independent Variables

- Socio-demographic variables like age, sex, educational status, income
- History of hypertension
- Family history of hypertension
- Alcohol abuse, cigarette smoking,
- Salt consumption
- Other co-morbid illnesses like diabetes, cardiac failure, or renal failure

### Data collection procedure

A closed-ended, structured questionnaire was used to collect data. Information on the patient’s details, clinical staging of the disease at the time of diagnosis, the duration of treatment, drug details, and the results of investigations performed were collected using a data collection format. It was collected by the principal investigator and four other medical interns working in the department.

### Data Quality Control

To ensure the completeness, accuracy, and consistency of data collection, one hour of training was given to the data collectors.

Data editing was conducted daily by the principal investigator to check for accuracy, consistency, and completeness. Data were entered and cleaned by the principal investigator before analysis

### Data Processing and analysis

After collecting, cleaning, and checking the data for completeness by the principal investigator, it was analyzed using SPSS version 26 software, and the results were expressed using appropriate frequency distribution and cross-tabulation for the selected variables. Association between the independent and dependent variables were tested using the odds ratio, and a 95 percent confidence interval was used to measure the strength of the association between the independent and dependent variables.

### Ethical consideration

A proposal for the study format was submitted to the Jimma University Department of Medicine Office to get work approval for the study. Ethics committee of Jimma University gave ethical approval for the study. After approval, an official letter was obtained from the Department of Medicine at Jimma University to get permission from Adama Hospital Medical College.

Each respondent was informed about the objective, purpose, and assurance of confidentiality. Before starting data collection, informed verbal consent was obtained from each subject. Patient identification, health care provider information, and drug product information were kept private, as their names were not displayed in the questionnaire or final report.

### Limitations of the study

This study used a prospective cross-sectional survey obtained directly from the patient and secondary data from a single hospital. The result may not be representative of the national or regional picture as it is done in one hospital involving a referral hospital population with a relatively well-organized surgical ward in the region. The diagnosis and patient condition were taken as recorded on the clinical records. Due to costs, some confirmatory investigations, such as a CT scan and cardiac biomarkers, were not available in all cases.

In some cases, it was difficult to determine whether the end-organ damage in this study was caused by the currently severely elevated blood pressure, an old previous lesion, or some other chronic disease.

### Operational definitions

**Prevalence** represents the total number of infections.
**Incidence** represents the number of new infections. It is a better indicator of the evolution of the epidemic, as it accounts only for new infections, while prevalence also accounts for deaths related to the infection.
**Hypertension** is defined by the average of two systolic blood pressures (SBP) between 130 and
139 and diastolic blood pressure (DBP) between 80 and 89 mmHg and/or current use of antihypertensive medications at the time of admission. (2)
**Severe Hypertension** is defined as average systolic blood pressure (SBP) ≥180 and or diastolic blood pressure (DBP) ≥120 mmHg.
**Hypertensive Emergency** is severe hypertension associated with end-organ damage. (2)
**Acute kidney injury** is currently defined by a rise in serum creatinine concentration from a baseline of at least 0.3 mg/dL within 48 hours or at least 50% higher than baseline within 1 week, or a reduction in urine output to below 0.5 mL/kg per hour for longer than 6 hours. (7)
**Acute myocardial infarction** was defined according to the previous World Health Organization’s criteria for acute, evolving, or recent myocardial infarction, which requires a combination of two of three characteristics: typical symptoms (i.e., chest discomfort), typical rise and gradual fall of troponin or more rapid rise and fall of CK-MB, and ECG changes indicative of ischemia (ST segment elevation or depression) involving the development of pathological Q-waves.
**Hypertensive encephalopathy** is severe hypertension with an alteration of mental status with no focal neurological deficits that resolved after lowering blood pressure.
**Acute pulmonary edema**: defined as the presence of dyspnea and bilateral basal crackles confirmed by a chest x-ray by a radiologist.
**Hypertensive Retinopathy** is an acute onset of blurred vision with retinal changes on funduscopy classified into mild, moderate, and severe

**Mild**: retinal arteriolar narrowing, wall thickening or opacification, and arteriovenous nicking (nipping).
**Moderate**: Hemorrhages, either flame or dot-shaped, cotton-wool spots, hard exudates, and microaneurysms.
**Severe**: Some or all above, as well as papilledema.
**Hypertensive Stroke** is severe hypertension with sudden onset of neurological deficits and confirmed by CT scan of the brain whether ischemic, hemorrhagic, or both.

## RESULTS

### Socio-demographic characteristics

During the study period, a total of 444 cases of hypertensive crisis were seen, of which 56.8% were men and the rest, 43.2%, were women. The most affected age groups are 66–75 years (42.8%) and 56–65 years (23.4%), followed by the age group >75 years, which accounts for 17.6% of the cases. 78.2% of the study subjects were from urban areas, and the rest, 21.8%, were from rural areas. (Table 1)

**Table 1:** Age, sex, and address of patients with hypertensive crisis presented to the EOPD at AHMC, Adama, Oromia, Ethiopia, from January 01 to August 31, 2021 G.C.

| Variables |  | Frequency | Percent |
| --- | --- | --- | --- |
| Age | <45 year | 15 | 3.8 |
|  | 45-55 year | 67 | 15.1 |
|  | 56-65 year | 104 | 23.4 |
|  | 66-75 year | 190 | 42.8 |
|  | >75 year | 78 | 17.6 |
|  | **Total** | 444 | 100.0 |
| Sex | Male | 252 | 56.8 |
|  | Female | 192 | 43.2 |
|  | **Total** | 444 | 100.0 |
| Place of Residency | Rural | 97 | 21.8 |
|  | Urban | 347 | 78.2 |
|  | **Total** | 444 | 100.0 |

Of the study subjects, 28.4% can read and write, followed by 25.7% who have attended primary school. (Figure 1) Around 45.9% of the patients have an average income per month of 3,880–15,135 birr, followed by 25.7% who earn 3,880 birr per month. (Figure 2)

**Figure 1:**
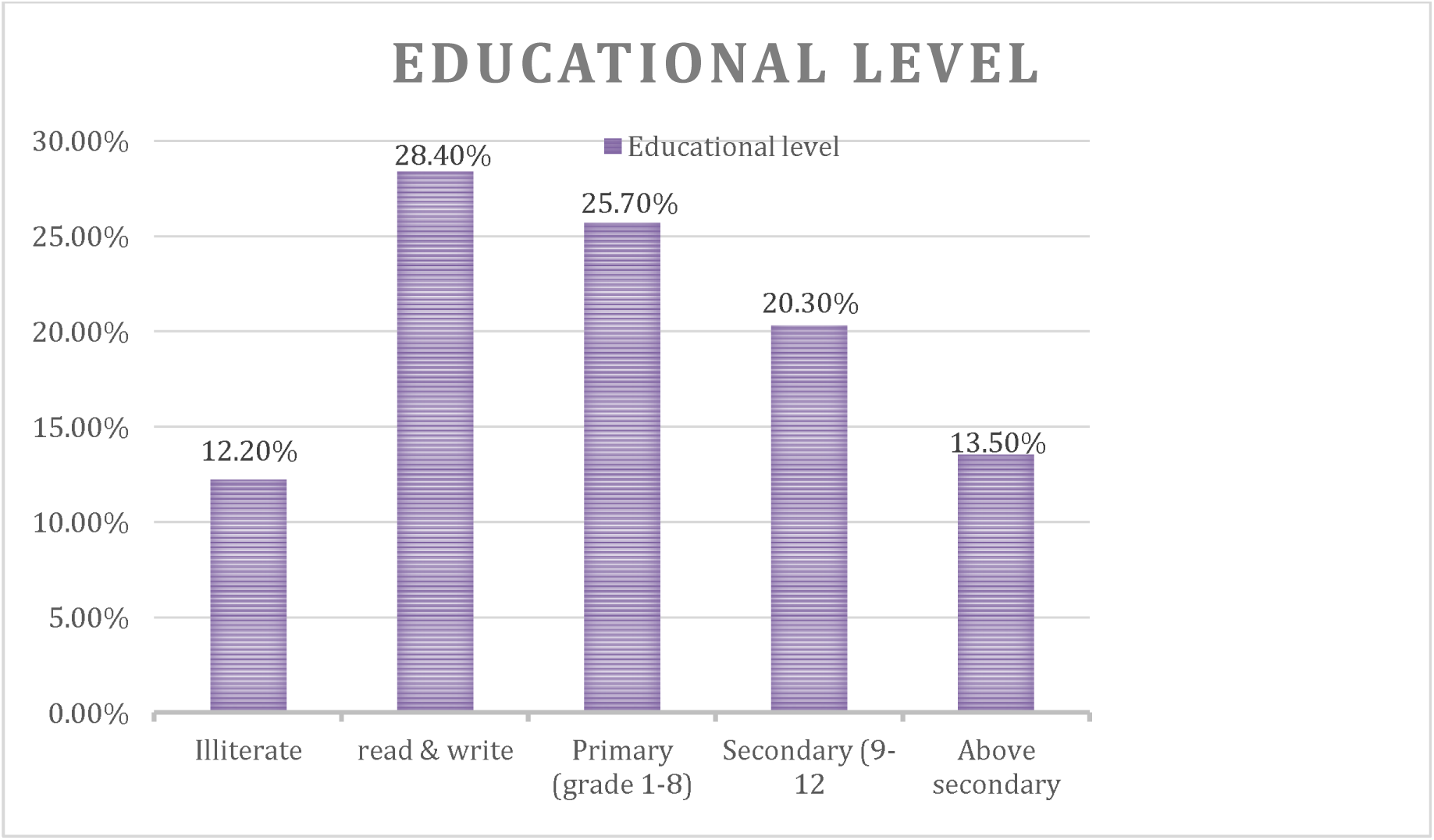
Educational Level of Hypertensive Crisis Patients Presented to EOPD at AHMC, Adama, Oromia, Ethiopia, from January 01 to August 31, 2021 G.C.

**Figure 2:**
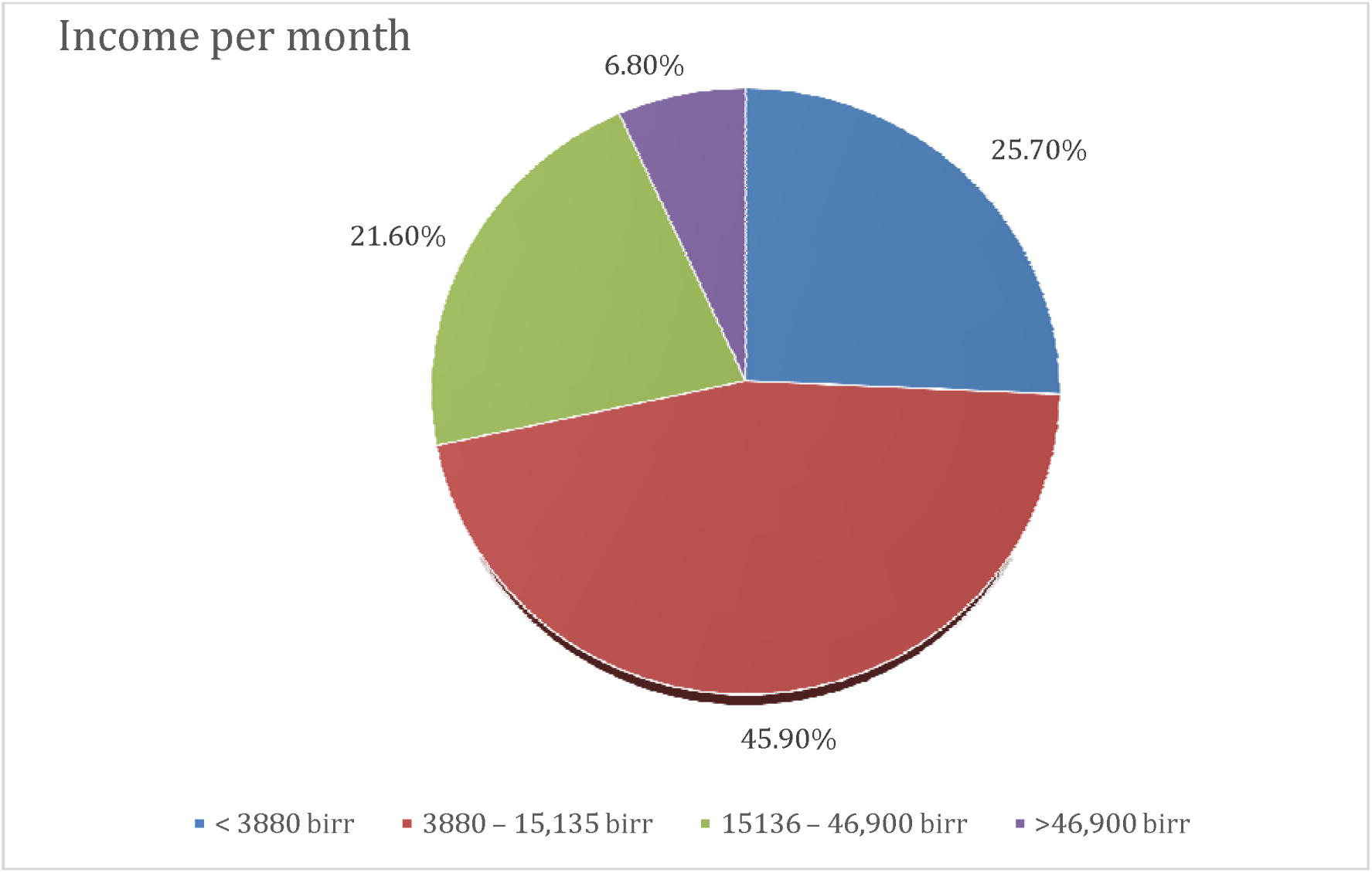
Income per month of patients with hypertensive crisis presented with EOPD at AHMC, Adama, Oromia, Ethiopia, from January 01 to August 31, 2021 G.C

### Clinical status of patients with a hypertensive emergency at presentation

At presentation, 91.0% of the study participants were known hypertensive patients, while 9.0% had just received a diagnosis. Of the known hypertensive patients, the majority (33.8%) were known to have been hypertensive for 5–10 years. Looking at the status of drug adherence, 48.6% were found to be adherent and 234 (52.7%) were on follow-up. (Table 2)

**Table 2:**
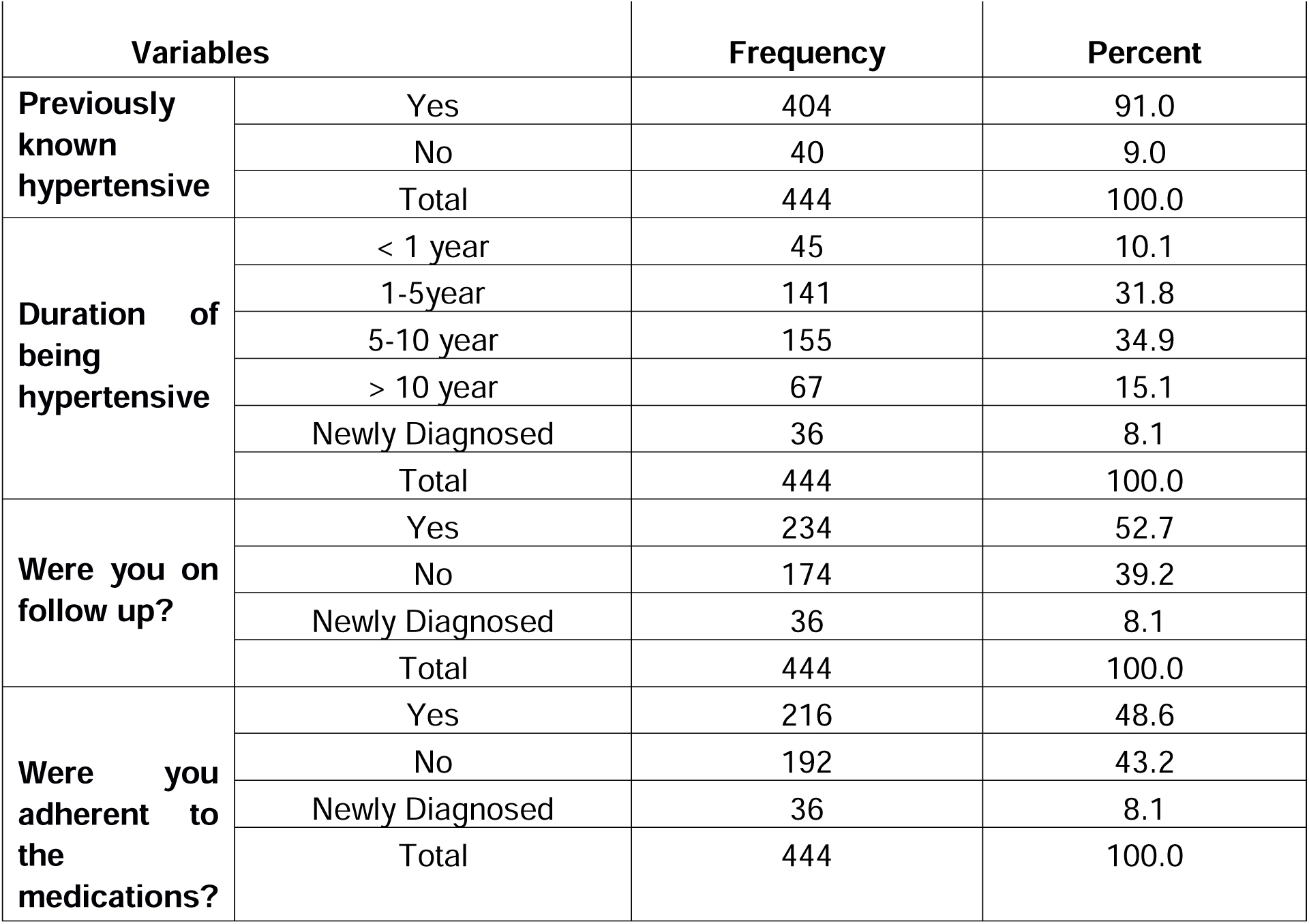
History of patients with hypertensive crisis presented to EOPD at AHMC, Adama, Oromia, Ethiopia, from January 01 to August 31, 2021 G.C.

Lack of knowledge was a major factor in 70 (40.2%) cases of those who were not on follow-up, followed by being too far from the health center and negligence, which each accounted for 34 (19.5%). Moreover, among those who do not take their medication, the main reasons are cost and a sense of well-being, which account for 28.1% of the total. (Table 3)

**Table 3:** Reasons for not being on follow-up of patients with hypertensive crisis presented to EOPD at AHMC, Adama, Oromia, Ethiopia, from January 01 to August 31, 2021 G.C.

| Variables |  | Frequency | Percent |
| --- | --- | --- | --- |
| If you are not on follow up, why? | Lack of knowledge | 70 | 40.2 |
|  | far from health center | 34 | 19.5 |
|  | Negligence | 34 | 19.5 |
|  | feeling of being well | 36 | 20.7 |
|  | Total | 174 | 100.0 |
| If you are not adherent, why? | Lack of knowledge | 34 | 17.7 |
|  | Cost | 54 | 28.1 |
|  | Negligence | 25 | 13.1 |
|  | Fear of side effects | 25 | 13.1 |
|  | feeling of being well | 54 | 28.1 |
|  | Total | 192 | 100.0 |

Most of the study subjects complained of headaches (43.9%), body weakness (27.7%), incidental findings on follow-up (12.0%), and loss of consciousness (8.1%). The duration of the complaint in most of the cases was less than a day (36.5%), followed by 24 to 72 hours (27.2%). (Table 4)

**Table 4:** Current complaints of patients with hypertensive crisis presented to the EOPD at AHMC, Adama, Oromia, Ethiopia, from January 01 to August 31, 2021 G.C.

| Variables |  | Frequency | Percent |
| --- | --- | --- | --- |
| Current Presenting Complaint | Headache | 195 | 43.9 |
|  | Body weakness | 123 | 27.7 |
|  | Loss of consciousness | 36 | 8.1 |
|  | Abnormal body movement | 24 | 5.4 |
|  | Impaired vision | 13 | 2.9 |
|  | Incidental on follow up | 53 | 12.0 |
|  | Total | 444 | 100.0 |
| Duration of compliant | < 24 hour | 162 | 36.5 |
|  | 24 to 72 hour | 121 | 27.2 |
|  | 3 to 7 days | 65 | 14.7 |
|  | 1 to 2 weeks | 30 | 6.8 |
|  | > 2 weeks | 10 | 2.3 |
|  | incidental finding | 56 | 12.6 |
|  | Total | 444 | 100.0 |

### Factors associated with Hypertensive Emergencies

Regarding the risk factors of patients with hypertensive crisis who were seen in the emergency room, salt consumption accounts for 52.7%, alcohol consumption is at 36.5%, family history of hypertension is at 32.4%, chewing tobacco is at 32.4%, and there is a prior history of severe HTN range. (Figure 3)

**Figure 3:**
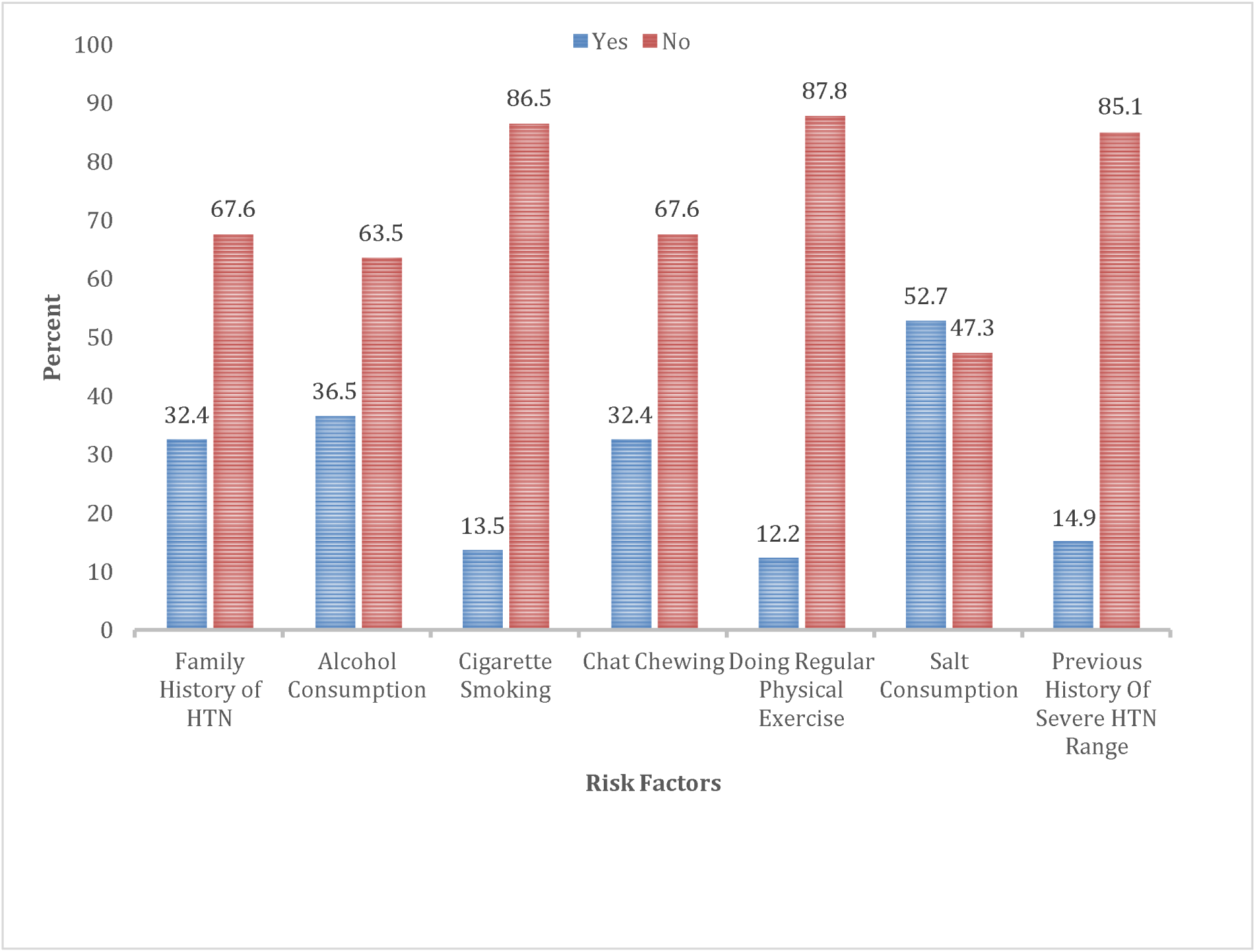
Risk factors of patients with hypertensive crisis presented to EOPD at AHMC, Adama, Oromia, Ethiopia, from January 01 to August 31, 2021 G.C.

Age was significantly associated with severe hypertension in the univariate analysis using the Pearson chi-square test of socio-demographic variables. The majority of patients with BP > 200/100 mmHg were > 56 years old, with the proportion increasing as age advances compared to those between 45 and 55 (P-value = 0.004). (Figure 4)

**Figure 4:**
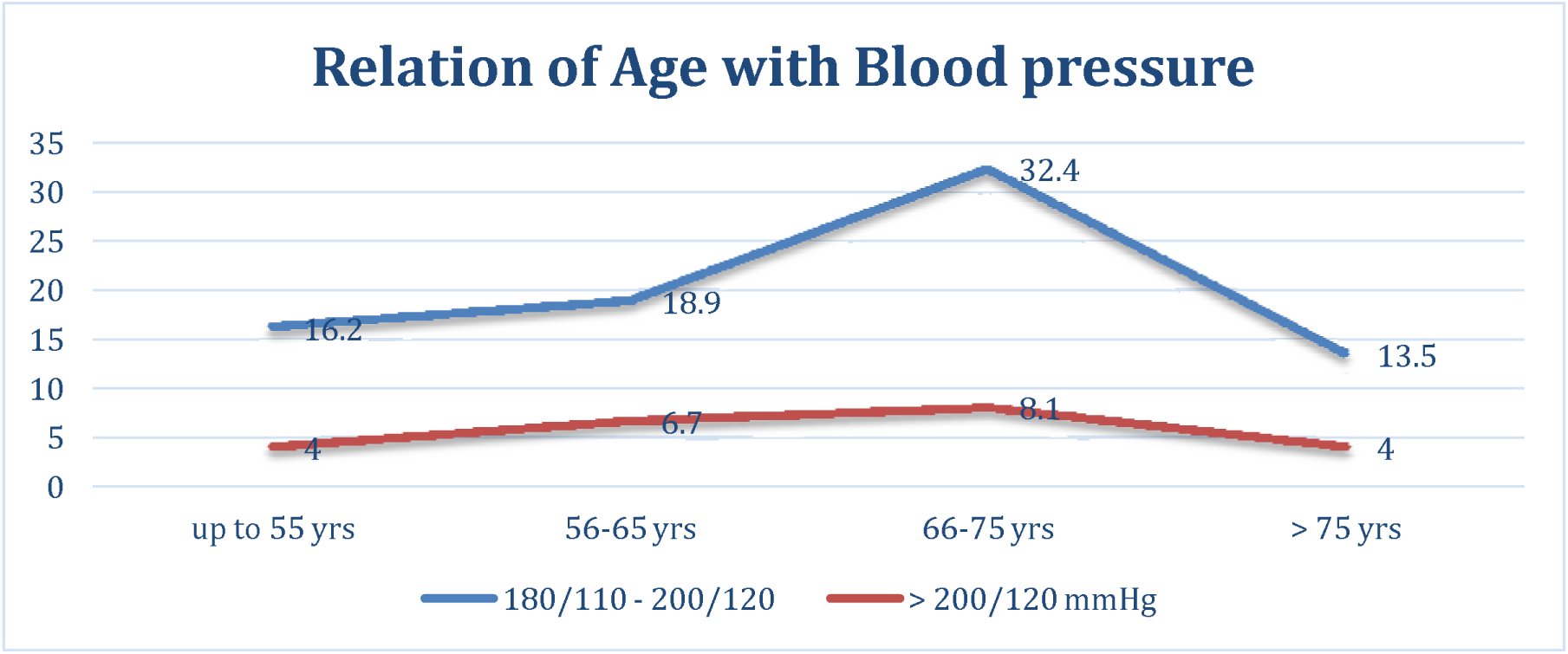
Relationship of age and blood pressure among patients with hypertensive emergencies presented to the EOPD at AHMC, Adama, Oromia, Ethiopia, from January 01 to August 31, 2021 G.C.

Similarly, when compared with those from rural areas, those from urban areas are more likely to present with severe hypertension than those from rural areas (P = 0.01). (Figure 5)

**Figure 5:**
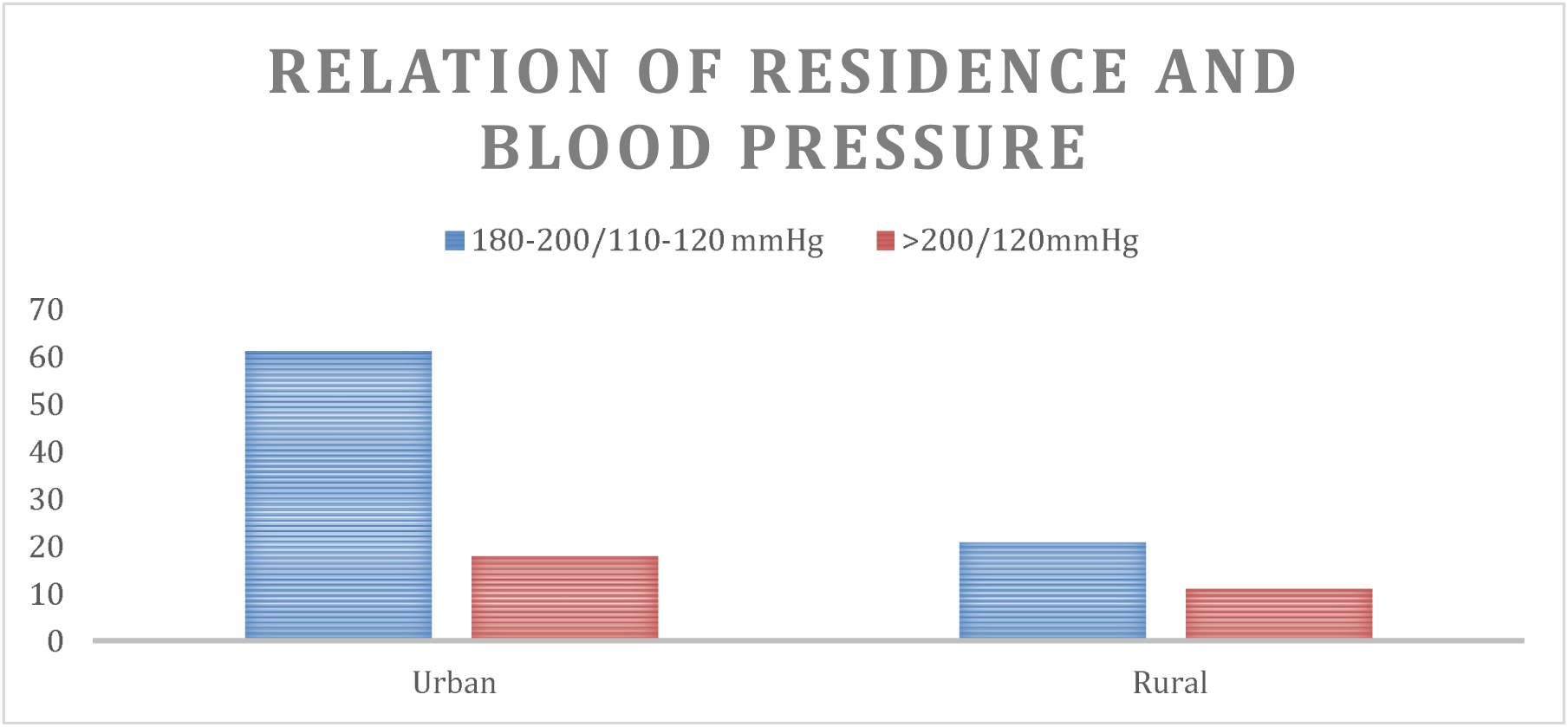
Relationship of residence and hypertension among patients with hypertensive emergencies presented to the EOPD at AHMC, Adama, Oromia, Ethiopia, from January 01 to August 31, 2021 G.C.

Associated comorbid illness was found among patients with hypertensive crises presenting to the emergency department, including diabetes mellitus in 108 (24.3%) of the cases, CHF in 30 (6.8%), renal disease in 24 (4.1%), and 6 (1.4%) of them had DM and CHF, and another 6 (1.4%) had DM and renal failure simultaneously, but there were no other identified comorbid illnesses in the majority of the cases (258, or 58.1%). (Figure 6)

**Figure 6:**
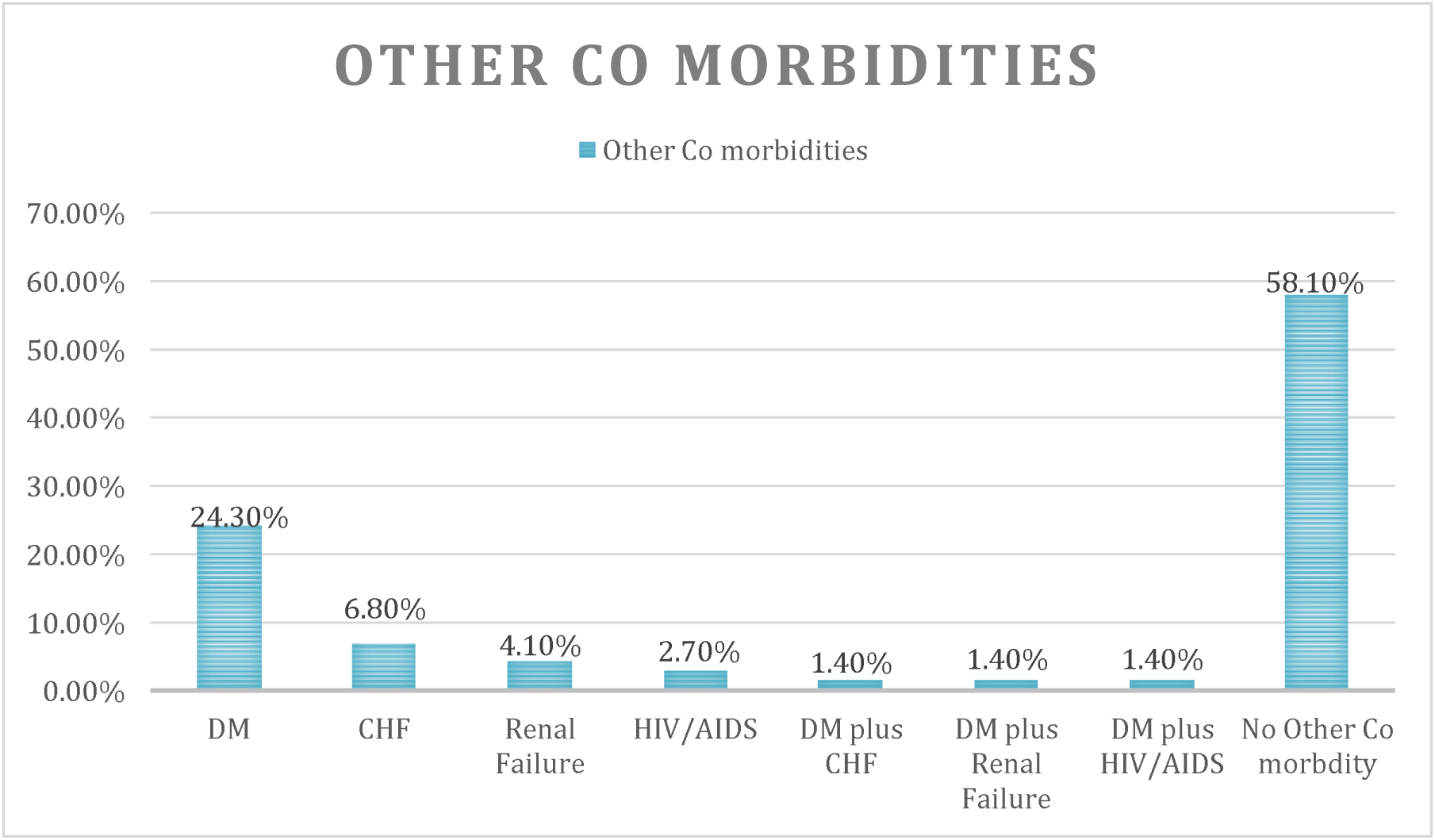
Other comorbidities of patients with hypertensive crisis presented to EOPD at AHMC, Adama, Oromia, Ethiopia, from January 01 to August 31, 2021 G.C.

### Physical, laboratory, and radiologic findings among patients with hypertensive crisis

On physical examination, the blood pressure of patients with hypertensive emergencies at initial presentation were SBP in the range of 180–190 mmHg and DBP of 110–115 mmHg in 222 (50.0%), followed by SBP of 191-200 mmHg and 116–120 mmHg in 137 (30.9%) of the cases, while others had SBP of greater than 220 and diastolic of above 130 in 24 (5.4%). In terms of BMI, 199 (44.8%) of the study population had a BMI between 18.5-24.9 kg/m^2^, with 107 (24.1%) having a BMI between 25-29.9 kg/m^2^. (Table 5)

**Table 5:** Physical findings of patients with hypertensive crisis presented to EOPD at AHMC, Adama, Oromia, Ethiopia, from January 01 to August 31, 2021 G.C.

| Variables |  | Frequency | Percent |
| --- | --- | --- | --- |
| **Blood pressure at presentation** | 180/110 - 190/115 | 222 | 50.0 |
|  | 191/116 - 200/120 | 137 | 30.9 |
|  | 201/121 - 220/130 | 61 | 13.7 |
|  | >220/130 | 24 | 5.4 |
|  | Total | 444 | 100.0 |
| **BMI** | <18.5 Kg/m<sup>2</sup> | 30 | 6.8 |
|  | 18.5 - 24.9 Kg/m<sup>2</sup> | 199 | 44.8 |
|  | 25 - 29.9 Kg/m<sup>2</sup> | 107 | 24.1 |
|  | 30 - 34.9 Kg/m<sup>2</sup> | 66 | 14.9 |
|  | 35 - 39.9 Kg/m<sup>2</sup> | 30 | 6.8 |
|  | >40 Kg/m<sup>2</sup> | 12 | 2.7 |
|  | Total | 444 | 100.0 |
| **Cardiac finding** | Displaced apex | 30 | 6.8 |
|  | No finding | 414 | 93.2 |
|  | Total | 444 | 100.0 |
| **Neurologic finding (GCS)** | 15/15 | 270 | 60.8 |
|  | 13 – 14 | 54 | 12.2 |
|  | 8 – 12 | 84 | 18.9 |
|  | <8 | 36 | 8.1 |
|  | Total | 444 | 100.0 |

Regarding the neurologic examination, the majority of patients 270 (60.8%) had a GCS of 15/15, while 54 (18.9%) had a GCS of 8–12 and 36 (8.1%) had a GCS of less than 8. On motor examination, 66 (14.9%) had hemiparesis, followed by 36 (8.1%) with hemiplegia. In 306 (68.9%) of the patients, there were no findings. (Figure 7)

**Figure 7:**
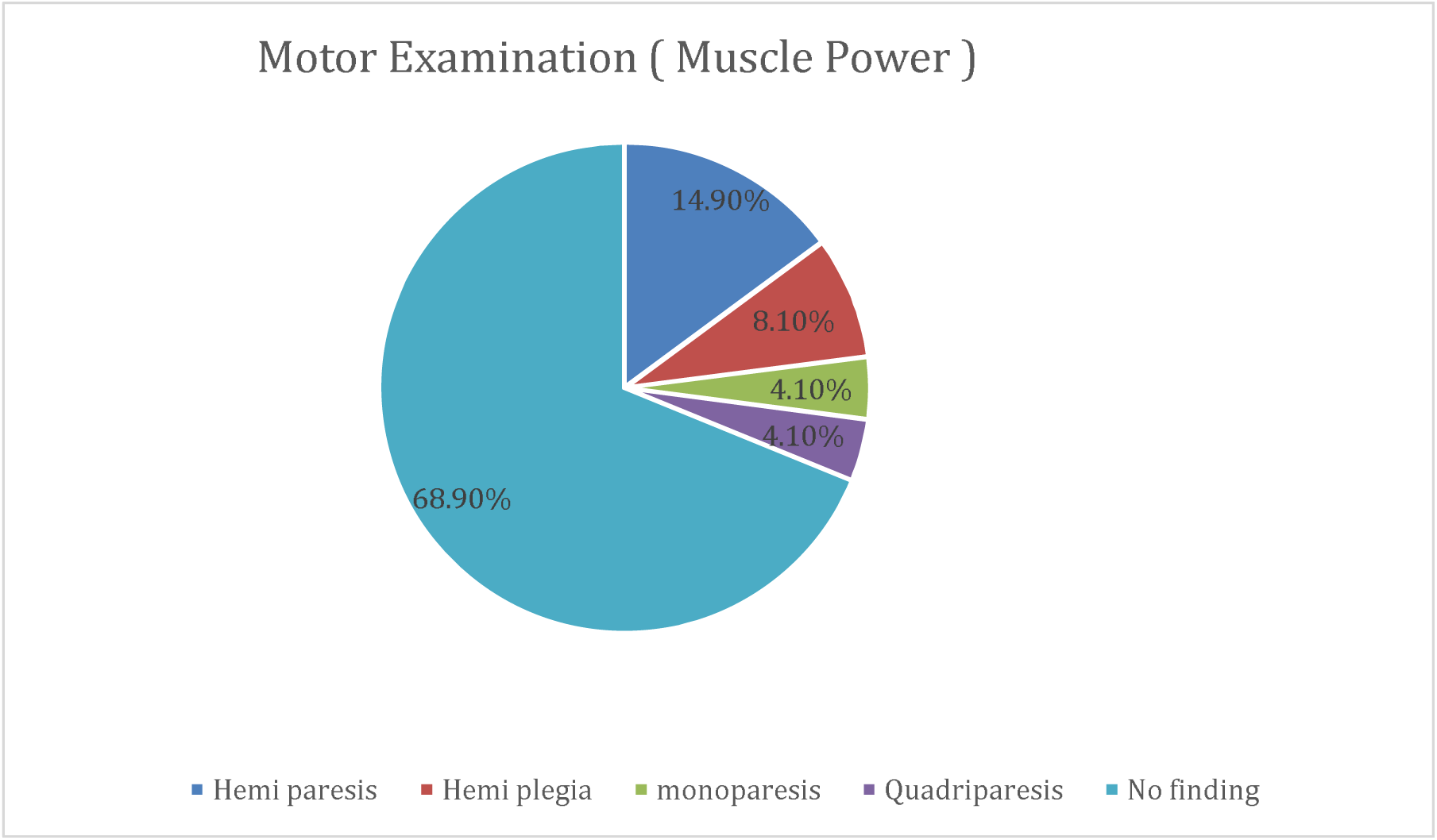
Motor examination (Muscle power) of patients with hypertensive crisis presented to EOPD at AHMC, Adama, Oromia, Ethiopia, from January 01 to August 31, 2021 G.C

On laboratory evaluation, the random blood sugar values were as follows: In more than half of the patients (74.5%), the random blood sugar values were in the range of 70–110 mg/dl; in 13.3% of the patients, the RBS was 111–200 mg/dl; and in 5.4% of the patients, the RBS was >200 mg/dl.

On serologic examination, serum creatinine values were in the range of 0.7–1.2 mg/dl in 366 (82.4%), 1.2-2 mg/dl in 53 (11.9%), and in 25 (5.6%) of the cases, serum creatinine was above 2 mg/dl. (Table 6)

**Table 6:** Laboratory findings of patients with hypertensive crisis presented to EOPD at AHMC, Adama, Oromia, Ethiopia, from January 01 to August 31, 2021 G.C.

| Variables |  | Frequency | Percent |
| --- | --- | --- | --- |
| FBS/RBS | <70 mg/dl | 30 | 6.8 |
|  | 70 - 110mg/dl ( Normal) | 331 | 74.5 |
|  | 111 - 200 mg/dl | 59 | 13.3 |
|  | >200 mg/dl | 24 | 5.4 |
|  | Total | 444 | 100.0 |
| Serum Creatinine | 0.7 - 1.2 mg/dl (normal) | 366 | 82.4 |
|  | 1.3 - 2.0 mg/dl | 53 | 11.9 |
|  | >2.0 mg/dl | 25 | 5.6 |
|  | Total | 444 | 100.0 |
| **Alanine Transaminase** | 0 - 40 U/L (normal) | 341 | 76.8 |
|  | 41 - 100 U/L | 36 | 8.1 |
|  | 100 - 200U/L | 24 | 5.4 |
|  | >200 U/L | 43 | 9.7 |
|  | Total | 444 | 100.0 |
| **Aspartate Transaminase** | 0 - 40 U/L (normal) | 347 | 78.1 |
|  | 41 - 100 U/L | 30 | 6.8 |
|  | 100 - 200U/L | 37 | 8.3 |
|  | >200 U/L | 30 | 6.8 |
|  | Total | 444 | 100.0 |
| **Triglycerides** | 0 - 150 mg/dl (Normal) | 360 | 81.1 |
|  | 150 - 200 mg/dl | 40 | 9.0 |
|  | >300 mg/dl | 44 | 10.0 |
|  | Total | 444 | 100.0 |
| **Cholesterol** | 0 - 200 mg/dl (Normal) | 354 | 79.7 |
|  | 200 - 300 mg/dl | 49 | 11.0 |
|  | >250 mg/dl | 41 | 9.2 |
|  | Total | 444 | 100.0 |

Most of them had normal chest x-ray findings on radiographic examination: 252 (51.8%) had normal chest x-ray findings and 60 (13.6%) had abnormal chest x-ray findings. ECGs were performed on 414 (93.2%) of the patients, and the ECGs were normal in 378 (85.1%), with only 12 (2.7%) of the patients showing STEMI. A CT scan of the head was performed on 204 (45.99%) of the patients, the majority of whom had a hemorrhagic stroke (12.6%). A CT scan of the head was not performed on 54.1% of the patients. (Table 7)

**Table 7:**
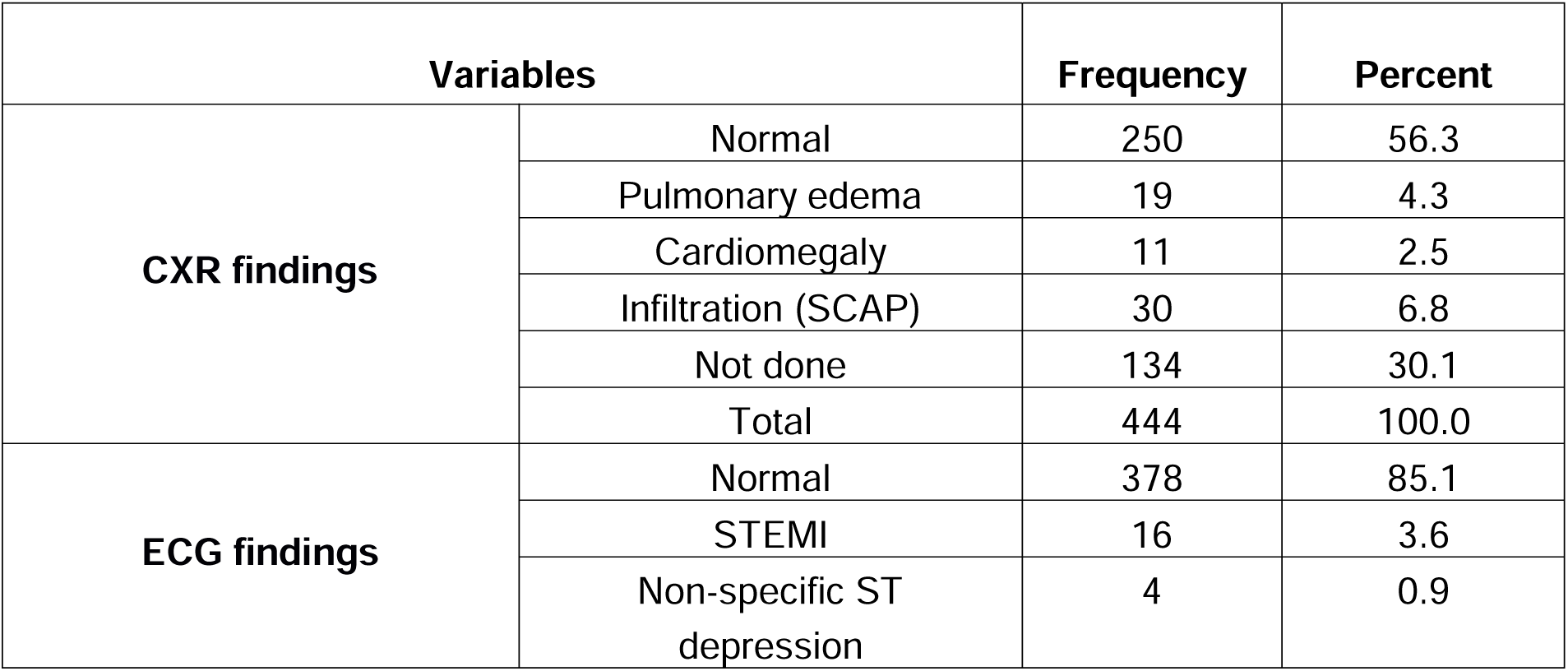
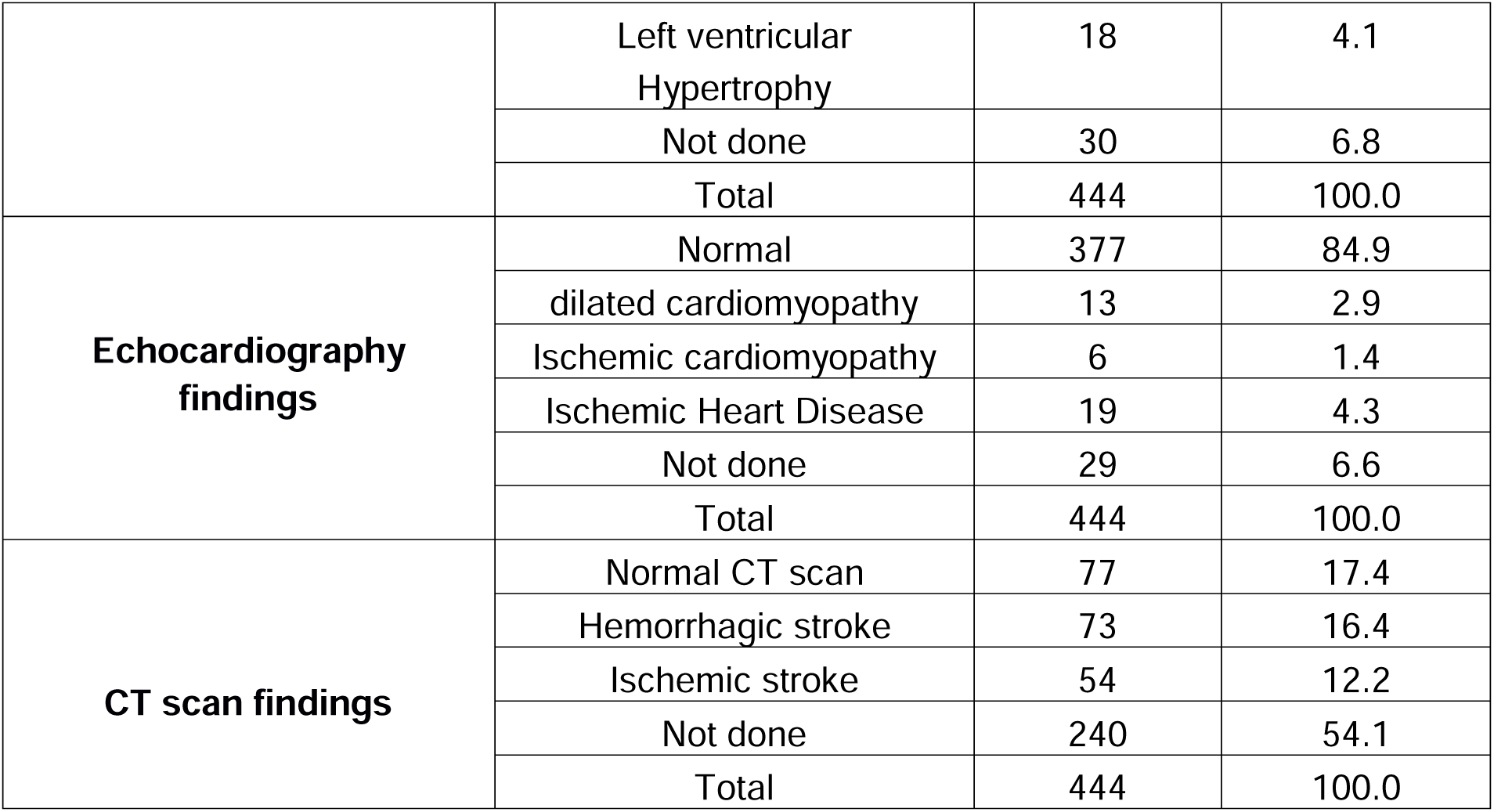
Radiologic findings of patients with hypertensive crisis presented to EOPD at AHMC, Adama, Oromia, Ethiopia, from January 01 to August 31, 2021 G.C.

After presenting to the emergency room, 12 (16.2%) patients are discharged within 24 hours. 27 (36.5%) patients stayed for 24 to 72 hours (about 3 days), and 22 (29.7%) stayed for 72 to 6 days. (Figure 8)

**Figure 8:**
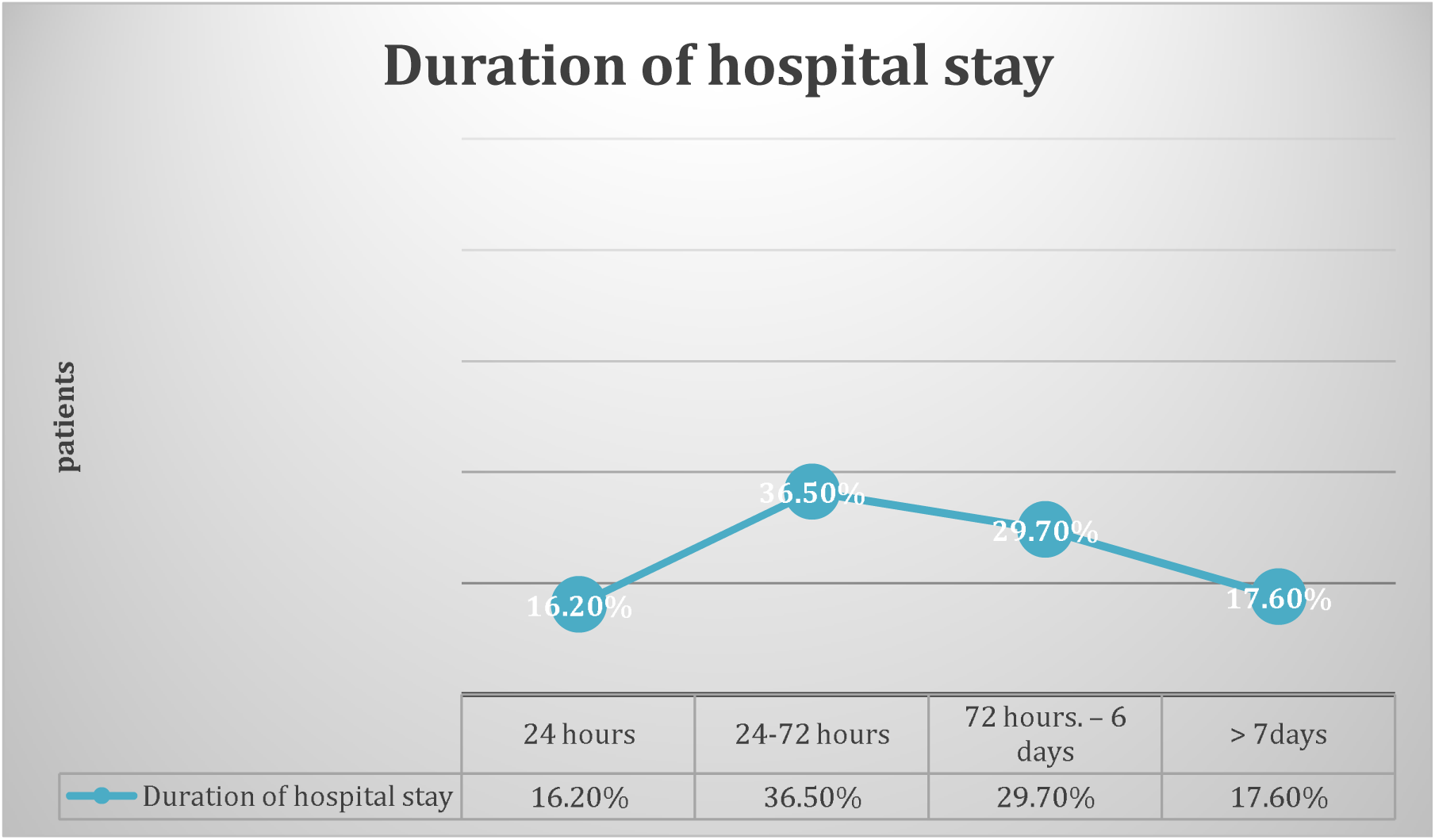
Duration of hospital stay of patients with hypertensive crisis presented to EOPD at AHMC, Adama, Oromia, Ethiopia, from January 01 to August 31, 2021 G.C.

## DISCUSSION

### Factors associated with hypertensive emergencies

In this study, 444 patients were enrolled, with men accounting for more than half (56.8%) of those diagnosed with hypertensive crisis, which is comparable to a study conducted in Brazil in 2000, which found that 55.3% were men and 44.7% were women. (11)

Moreover in this analysis, an increase in age above 55 years was found to be a significant risk factor for hypertensive emergencies (P = 0.004), as shown in the univariate analysis. These findings were similar to those of studies done in the USA, where old age was one of the independent predictors of poorly controlled hypertension (12). Similar results were obtained from a study conducted in Nigeria, where it was discovered that among black hypertensive patients who sought primary medical care, old age was most frequently linked to target organ damage (13).

Most of the patients (91.0%) with hypertensive emergencies were known hypertensive patients, and 40 (9.0%) of them were newly diagnosed. Only 39 (52.7%) were on follow-up, and 36 (48.6%) were found to be adherent; a major reason was a lack of knowledge, accounting for 41.4% of cases, and a sense of improvement in 28.1% of cases, particularly in rural areas where being far from the health center and cost were also factors. These studies are comparable with a prospective study done on patients in the cardiac emergency ward of Imam Raza Hospital, which showed that 75% of them had discontinued taking their medication for a long time, with the most common reason being a feeling of improvement. (14)

Salt consumption accounts for 234 (52.7%) of the risk factors for patients with hypertensive crises who present to emergency department in our study, followed by alcohol consumption (36.5%). Moreover, when we see the BMI of our study population, 107 (or 24.1%) of them are between 25 and 29.9 kg/m2. These are comparable with a population-based, cross-sectional survey done in Addis Ababa, which showed that about 20% of males and 38% of females were overweight (body mass index ≥ 25 kg/m^2^). (15)

Furthermore, in the univariable analysis, our study found that a history of diabetes mellitus was a significant predictor of hypertensive emergencies (P = 0.002). This is similar to a study done in Brazil in which diabetes mellitus was found in 20% of patients and was found to be a statistically significant factor for the development of hypertensive emergencies (16). Moreover, a study in Ghana showed that participants diagnosed with diabetes had increased odds of developing target organ damage compared to those with no diagnosis of diabetes (17).

Our study also found that those from urban areas are significantly more associated with hypertensive emergencies and complications than those from rural ones. This can be explained by the current adoption of western culture, including diet changes, smoking, obesity, and life changes associated with urbanization, all of which are associated with cardiovascular complications. However, a study conducted in the Congo by Ellenga et al. revealed that hypertensive emergencies were significantly associated with low socioeconomic status (18).

### Complications of hypertensive emergency

In our study, stroke was a major complication, accounting for 28.6%. The number of emergencies from hemorrhagic stroke was 27.1%, which was higher than in previous studies. Stroke was the most prevalent disease, accounting for 50% of the cases (19) in the study from the Democratic Republic of the Congo, whereas in the Italian study, stroke was responsible for 22% of the cases (6).

Furthermore, 8.2% of the patients had signs of heart failure contributing to cardiomegaly, left ventricular hypertrophy, and 4.1% had basal crepitation on chest examination. But in the Italian study, 121 patients (30.9%) had acute pulmonary edema.

In Brazil, too, acute pulmonary edema was one of the most common hypertensive emergencies (16). Our study has shown that 8.1% of patients with severe hypertension had renal dysfunction, which is lower than in the Congo, where renal failure was found in 13.1% of patients with severe hypertension (19), and again lower than in Italy, where 9.9% had acute renal failure (6). These showed that hypertensive kidney diseases are undiagnosed because of the limited facilities in our setting.

## Conclusions

Hypertensive crises are one of the most common causes of admission among patients who visit the Emergency Outpatient Department (EOPD) and are associated with high rates of complications. Most of the patients were males between the ages of 66 and 75. More than half of the patients were from urban areas.

Factors commonly associated with hypertensive emergencies include age above 55 and the patients’ place of residence. Most of the study subjects at presentation were known hypertensive patients; of this majority, they had been known to be hypertensive for 5–10 years. Less than half of the patients were found to be adherents, the major reason being a lack of knowledge.

Patients with hypertensive crises who presented to the emergency department had associated comorbid illnesses, and diabetes mellitus was found in a quarter of them.

The most common forms of hypertensive crisis include stroke, cardiac dysfunction, and renal dysfunction.

### Recommendations

The government should implement more emphasis on population-wide hypertension prevention and management, which is necessary. The potential complications of hypertension should be explained to patients. To provide hospitals with the necessary baseline investigations and to enhance the outcomes of hypertensive patients, infrastructure and capacity building are required.

Healthcare providers should educate patients on the importance of consistent use of medication on discharge to reduce the case fatality rate post discharge. There is a need to educate the community on the importance of lifestyle modifications such as dietary modification and exercise, to reduce the risk of hypertension and to develop a habit of checking their blood pressure for early detection and treatment to avoid the complications of hypertension, which are fatal. Patients should be evaluated and managed for possible records, including initial and follow-up laboratory investigation results.

## Data Availability

All data produced in the present study are available upon reasonable request to the authors

## Abbreviations

ACC: American College of Cardiology
AHA: American Heart Association
AHMC: Adama Hospital Medical College
MAP: Mean Arterial Pressure
BMI: Body Mass Index
BP: Blood Pressure
CHD: coronary heart disease
CHF: Congestive heart failure
CKD: chronic kidney disease
CT: Computed Tomography
CXR: Chest X-Ray
DBP: Diastolic blood pressure
ECG: Electrocardiography
EOPD: Emergency Outpatient Department
HIV: Human Immunodeficiency Virus
HTN: Hypertension
ISH: International society of hypertension
JNC: Joint National Committee
NCHS: National community health statistics
RBS: Random Blood Sugar
SSA: Sub-Saharan Africa
SBP: Systolic blood pressure
WHO: World Health Organization

## Declarations

### Ethical approval and consent to participate

Data collection was initiated after an ethical approval letter was obtained from the Faculty of Medical Sciences of Jimma University (IRBJU/20/2021) and Adama Hospital Medical college allowed the data collection process which gave the investigator a grant as a waiver for patients’ consent. The data is anonymous with absolute confidentiality of the patients.

### Consent for publication

Not applicable.

### Availability of data and materials

Data is submitted.

### Author’s contribution

The study’s conception, tool development, proposal development, logistics, data management, data collection processes, design, analysis, and manuscript assembly have all involved ATA, YTK, and BDM. The manuscript has been reviewed and approved by all authors.

## Acknowledgement

We are grateful to Jimma University’s School of Medicine for allowing us to undertake this research. We thank the staff at Adama Hospital Medical College for their assistance with data collection.

